# The effect of minimum unit pricing for alcohol on prescriptions for treatment of alcohol dependence: a controlled Interrupted Time Series analysis

**DOI:** 10.1101/2022.12.08.22283246

**Authors:** Francesco Manca, Lisong Zhang, Niamh Fitzgerald, Daniel Mackay, Andrew McAuley, Clare Sharp, Jim Lewsey

## Abstract

In 2018, Scotland introduced a minimum unit price (MUP) for alcohol to reduce alcohol-related harms. We aimed to study the association between MUP introduction and the volume of prescriptions to treat alcohol dependence, and volume of new patients receiving such prescriptions. We also examined whether effects varied across different socioeconomic groups. A controlled interrupted time series was used to examine variations of our two outcomes. The same prescriptions in England and prescriptions for methadone in Scotland were used as controls. There was no evidence of an association between MUP implementation and the volume of prescriptions for alcohol dependence (immediate change: 2.74%, 95% CI: -0.068 0.014; slope change: 0% 95%CI: -0.001 0.000). A small, significant increase in slope in number of new patients receiving prescriptions was observed (0.2% 95%CI: 0.001 0.003). However, no significant results were confirmed after robustness checks. We found also no variation across different socioeconomic groups.

## Introduction

Alcohol is one of the leading risk factors for premature death and disability worldwide [1]. In the United Kingdom, the incidence of alcohol-related harm is above the world average, being the fifth-ranked cause of death and serious illness [2]. Evidence shows that since the mid-1990s, alcohol sales per adult have been consistently higher in Scotland than in England & Wales [3], causing higher alcohol related harms as well as consumption [4]. Alcohol-related harm has also been shown to be more prevalent in certain sections of the population, widening health inequalities [5]. This is particularly evident in Scotland, where alcohol-related deaths are more than five times higher in the most socio-economically deprived areas compared to the least deprived areas [6].

The Scottish Government, recognising high levels of alcohol consumption as a major public health threat for its population, has implemented policies such as the Alcohol Act (restricting alcohol promotions within retail stores and banning quantity-based price discounts) in 2010 [7] and more restrictive drink-driving laws in 2014 [8]. Building on these earlier policies, and after a long legal battle, a minimum unit price (MUP) for alcohol came into effect in May 2018. MUP was intended to reduce alcohol consumption across the population, but to have greater effect on those who drink the most and favour cheaper products [9]. Specifically, MUP is a policy which sets a floor price of £0.50 ($0.57 or €0.58 – converted in September 2022) per UK unit of alcohol (one UK unit contains 8g of ethanol) and applies to all alcohol sold including in shops or bars/clubs. After 1st May 2018, alcohol products in Scotland could not legally be sold at any price equivalent to or below £0.50 per UK unit.

The introduction of MUP was supported by findings generated using the Sheffield Alcohol Policy Model (SAPM) [10], an epidemiological and econometric model that estimated the potential causal impact of the policy. SAPM estimated a direct reduction in the level of alcohol consumption due to MUP, which in turn would reduce related health harms and crime [11] and reduce inequalities. SAPM focuses on overall effects and broad sub-groups of the population including hazardous and harmful drinkers, but is not well suited to estimating the impact of MUP on people with more severe alcohol dependence, who are poorly represented in the data on which the model relies. The legislation introducing MUP is subject to a ‘sunset clause’ meaning that the policy will lapse if the Scottish Parliament does not vote for it to continue in 2023. To inform this parliamentary process, MUP is being evaluated by a suite of studies [12], some commissioned by the national agency for public health, and some funded separately. Studies published to date have shown mixed findings. First, there is strong evidence from separate research using different data sources that there has been an overall reduction in alcohol sales associate with MUP [13, 14]. However, in the early reporting on harms, no significant variations in crime and disorder [15] or attendances to emergency departments [16] were found after the introduction of MUP. Little is known about the effect of MUP on dependent drinkers. Early studies prior to the introduction of the policy suggested that the most dependent drinkers found it hard to understand that they could not ‘shop around’ for a better price, but some felt that they would cut down their alcohol intake under MUP [17]. A before- and after-MUP study of people drinking at harmful levels [18] found no clear evidence that level of alcohol consumption changed, nor severity of dependence. Finally, a qualitative study [19] found that introduction of MUP had little/no impact on people experiencing homelessness and the support services they use.

The primary aim of this study was to investigate the effect of the introduction of MUP on the level of prescriptions for the treatment of alcohol dependence and of new patients receiving such prescriptions. A secondary aim was to determine whether there was variation in any effect of MUP across different socio-economic groups.

## Methods

### Background

The National Institute for Health and Care Excellence (NICE) guideline for treatment of alcohol-use disorders [20] outlines that pharmacological intervention can be considered in combination with psychological interventions for treatment of alcohol dependence, and in particular recommends acamprosate, disulfiram, naltrexone and nalmefene [21]. Naltrexone is also used as a treatment for opiate dependence. As there was no way to distinguish naltrexone prescriptions to treat alcohol or opiate dependence in our dataset, we restricted the set of medications under study to acamprosate, disulfiram and nalmefene.

### Design

We used a controlled interrupted time–series (ITS) design to evaluate whether the introduction of MUP was associated with a change in the volume of prescriptions for treatment of alcohol dependence. To account for potential substitution effects between different medications, we assessed the impact of the legislation on the total volume of prescriptions for acamprosate, disulfiram and nalmefene combined (primary outcome measure). In addition, we used the number of new patients receiving prescriptions for treatment of alcohol dependence for the first time as a secondary outcome measure. As well as modelling for the entire population, we also ran ITS models for individuals residing in the most socio-economically deprived group (based on highest decile of Scottish Index of Multiple Deprivation [22]) and the remaining population.

To produce robust and reliable findings, controlled ITS designs are usually preferred to uncontrolled designs, as long as appropriate controls can be identified. The use of a control group that is equally affected by other potential factors happening in the same time period should minimise potential confounding [23]. Theoretically, using England, a location-based control group that neighbours Scotland, has the same UK government and culture and also influenced by NICE guidelines would have been ideal. However, England had a publicly available dataset with a smaller level of granularity (only aggregated monthly volumes) and only on prescriptions (not on patients). This meant only a descriptive comparison with England was possible. Therefore, as an alternative, we ran inferential comparisons on the same outcomes (both prescriptions and patients) using prescriptions in Scotland for methadone as a control outcome [23]. This was chosen as a drug for a different form of dependence –opioid – but not affected by MUP). Data on methadone prescriptions and patients were obtained from the same source as for alcohol dependence prescriptions. As recommended elsewhere [23], controls were chosen based on *a priori* evaluation of potential confounding events. To assess the appropriateness of the controls, we first compared pre-intervention trends descriptively. For the inferential comparison we also assessed whether the intervention and control time series followed a common pre-intervention trend [24]. Nevertheless, we were aware that the population identified by the control group in the inferential analyses (opioid dependent) refers to a population with often very different characteristics to individuals with alcohol dependence, and the prescriptions may therefore be affected by different external factors. Therefore, we performed an extensive range of sensitivity analyses both on the uncontrolled interrupted time series and the control group.

### Data

Data on prescriptions issued in Scotland were extracted from the Scottish national prescribing information system (PIS) for the period March 2014 to March 2020. PIS records all medicines prescribed and dispensed in the community in Scotland [25]. The PIS dataset is arranged by patient identifier and paid date (the date on which the prescription item is submitted for payment - which is always the last day of the month). Data are aggregated with daily frequency. Whereas this date of payment is always recorded, the date on which the prescription was issued (‘prescribed date’) or when the medication was dispensed by the pharmacy (‘dispensed date’) are not consistently recorded. Where not recorded, the prescribed and dispensed date default to be the same as the date of payment. Therefore, the last day of the month includes not only the drugs prescribed and dispensed on that day, but also all those paid that month for which no prescribed or dispensed date was recorded. As a result, there is a peak in prescription numbers in the data on the final day of every month, which sometimes exceeds by 8-10 times the daily average for that month. Conscious that an analysis grouping observations by month would have solved these issues but would have also reduced the number of observations after MUP introduction (23 data points), we smoothed the daily data by re-distributing the peak of prescriptions at the end of the month throughout the daily levels (see supplementary material). Data on English prescriptions were from the publicly available English prescribing dataset [26] and contained aggregated monthly prescriptions issued in England (with no information on patients).

### Descriptive analysis

We calculated the monthly volume of prescriptions and new patients receiving such prescriptions before and after MUP, both overall and by socio-economic deprivation groups. As there were 23 months available after MUP introduction (May 2018 – March 2020), to identify a potential difference in prescription levels excluding possible seasonal trends, we reported and compared values for the same months over three periods: two before the intervention to remove already present trends and possible influences of a national shortage of disulfiram [27, 28] which occurred between January and October 2017, and one after.

### Inferential analysis

We used inferential models with additive seasonal autoregressive integrated moving average errors as our main statistical approach. After testing the series for stationarity, candidate models were based on autocorrelation and partial autocorrelation plots of the data and adjusted based on the correlogram of the errors. Best-fitting models were then selected using the Akaike and Bayesian information criteria and error white noise assumption was assessed by Portmanteau’s test. To correct data for skewness, data were log transformed, this also allowed to interpret model covariates as percentage variation of the outcome variable. New patients receiving methadone, having three weeks over the study period with zero new patients, were not log transformed.

We estimated the size of the MUP effect by including a dummy variable in the regression assuming value 1 after MUP introduction and 0 before. A post intervention trend was also included in the regression to get an estimate of the continuing effect of MUP, that is, the slope of the change in successive time periods. Two additional dummy variables were added in the regression to account for recurrent or unusual events in the time series period. Specifically, they were, one at the end/beginning of every year to reflect the period when practitioners release a lower number of prescriptions, and one between January and October 2017 to consider the drop in prescriptions due to the national shortage of disulfiram.

### Sensitivity analyses

We performed sensitivity analyses to test the robustness of our results. Specifically, we used falsification tests simulating the intervention six months before or six months after the intervention.

As the drop in disulfiram prescriptions was relatively close to our intervention date, it may have affected our estimates whenever it could have affected physicians’ future prescription attitude (a steadiness of prescriptions below 2016 volumes just after the shortage and before MUP introduction could suggest so). Therefore, whenever MUP coefficients (level or slope changes) assumed statistically significant values, an additional change in slope variable just after the shortage in disulfiram was inserted to assess this likely spread and lagged effect of disulfiram shortage avoiding potential biases. The same model using shortage of disulfiram variables and removing MUP was also performed, comparing size and significance of coefficients.

For the groups satisfying common trend assumptions [24], the analysis on the difference between intervention and control was performed, and whenever MUP had statistically significant values, falsification tests were performed on the difference.

## Results

### Descriptive analysis

#### Prescriptions

Acamprosate and disulfiram were the most prescribed drugs for alcohol dependence, however, there was a greater relative difference between the volume of these two drugs in England compared to Scotland (see Figure 1b). Disulfiram prescriptions dropped in 2017 (Figure 1) due to a national shortage of supply from one of the main national wholesalers This generated an overall decrease in prescriptions for alcohol dependence. Scotland, having a relatively higher demand for disulfiram, was more affected than England. Overall, there has been a gradual increase in the volume of prescriptions between 2014 and 2020 (Figure 2, Table 1). During the national shortage of disulfiram (January-October 2017), prescriptions remained steady (+0.3%) compared to the same period the previous year. Ruling out the period affected by the dilsulfiram shortage, in the 21 months after the intervention there was a general increase in prescriptions (4.6%) compared to the same period from May 2014 to January 2016. On average, more than a fifth of prescriptions were for people residing in the most socio-economically deprived decile, and greater growth was registered for this group (12.7%). In England, prescriptions had an overall decreasing trend. The decrease decelerated in the last 21 months (−6.2%) compared to the previous two periods (−7.2%). The distribution in prescriptions across socio-economic deprivation deciles was similar to Scotland, however, prescriptions in the most socio-economically deprived groups decreased at a higher rate than in the rest of the population. Methadone prescriptions increased in the second time period and then decreased in the last one after MUP implementation, having an overall decline, mainly led by the most socio-economically deprived groups.

**Table 1.** Volume of prescriptions and number of new patients with an exclusive indication for alcohol dependence †.

|  | Intervention Group<br>(Scottish -alcohol dependence- prescriptions and patients ) |  | Location based control group<br>(England) | Control outcome<br>(Methadone) |  |
| --- | --- | --- | --- | --- | --- |
|  | Prescriptions | New patients | Prescriptions | Prescriptions | New patients |
| **Total population** |  |  |  |  |  |
| May 2014-Mar 2016 | 89738 | 6102 | 359818 | 568828 | 3284 |
| May2016-Mar2018 | 90040 (+0.3%) | 5031 (-23.7%) | 332487 (-7.6%) | 592210 (+4.1%) | 2974 (-9.4%) |
| May2018-Mar2020 | 93899 (+4.3%)*(+4.6%)** | 4656 (-7.5%)*(-23.7%)** | 311730 (-6.2%)*(-13.4%)** | 559731 (-5.5%)*(-1.6%)** | 2583 (-13.1%)*(-21.3%)** |
| **Most deprived decile\*\*\*** |  |  |  |  |  |
| May2014-Mar2016 | 20761 | 1301 | 87924 | 212948 | 875 |
| May2016-Mar2018 | 22359 (+11.1%) | 1099 (-4.3%) | 77166 (-12.2%) | 211915 (-0.0%) | 774 (-4.3%) |
| May2018-Mar2020 | 22691 (+1.5%)*(+12.7%)** | 1010 (-8.1%)*(-22.4%)** | 70869 (-8.2%)*(-19.4%)** | 191595 (-9.6%)*(-10%)** | 673 (-13%)*(-23.1%)** |
| **Rest of population\*\*\*** |  |  |  |  |  |
| May2014-Mar2016 | 69175 | 4750 | 221062 | 351039 | 2341 |
| May2016-Mar2018 | 67439 (-2.5%) | 3918 (-17.5%) | 215162 (-2.7%) | 377303 (7.5%) | 2135(-8.8%) |
| May2018-Mar2020 | 70973 (+5.2%)*(+2.5%)** | 3625 (-7.5%)*(-23.7%)** | 203978 (-5.2%)*(-7.8%)** | 365377(-3.2%)*(4.1%)** | 1857 (-13%)*(-20.1%)** |

†=The descriptive comparison was on both prescriptions and number of new patients for Scottish prescriptions in alcohol dependence and methadone and only on alcohol dependence prescriptions for English data.
\* Incremental percentage referring to May2016-March2018
\*\* Incremental percentage referring to May2014-March2016 to avoid the inclusion of shortage in disulfiram
\*\*\* Most deprived group and rest of population do not add up to overall population figures due to missing data in patients regarding socio-economic deprivation.

**Figure 1.**
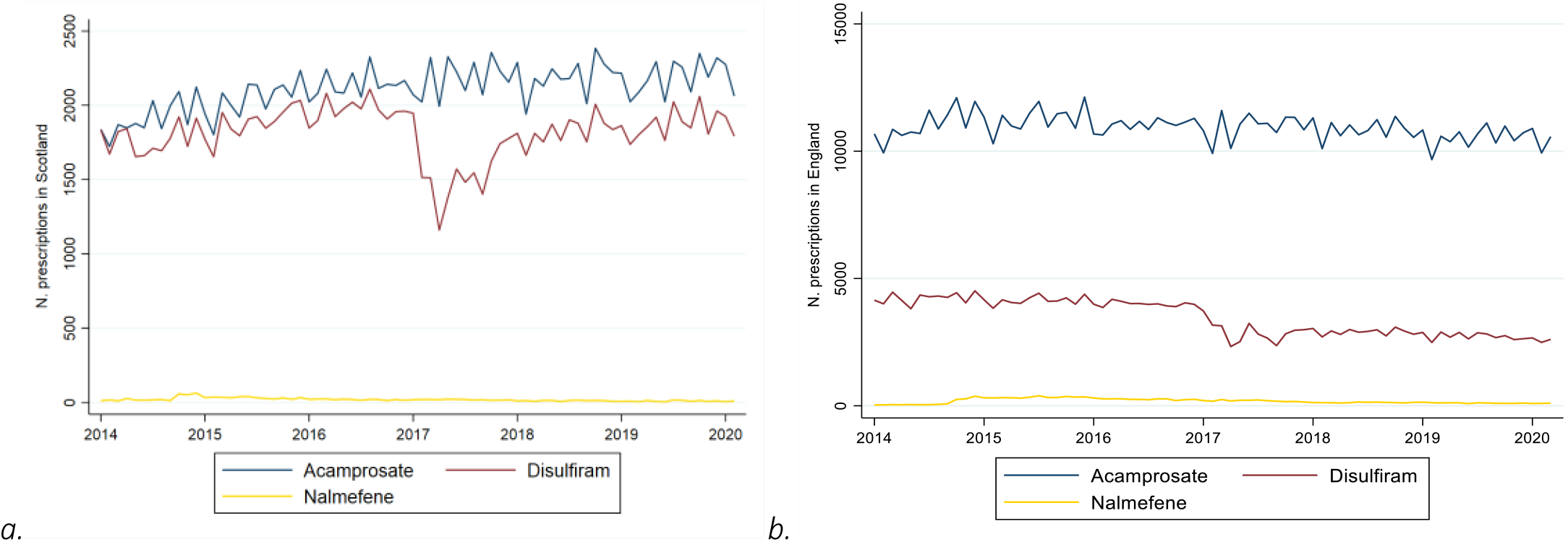
Prescriptions for alcohol dependence per month between 2014 and 2020 In Scotland (a) and England (b).

**Figure 2.**
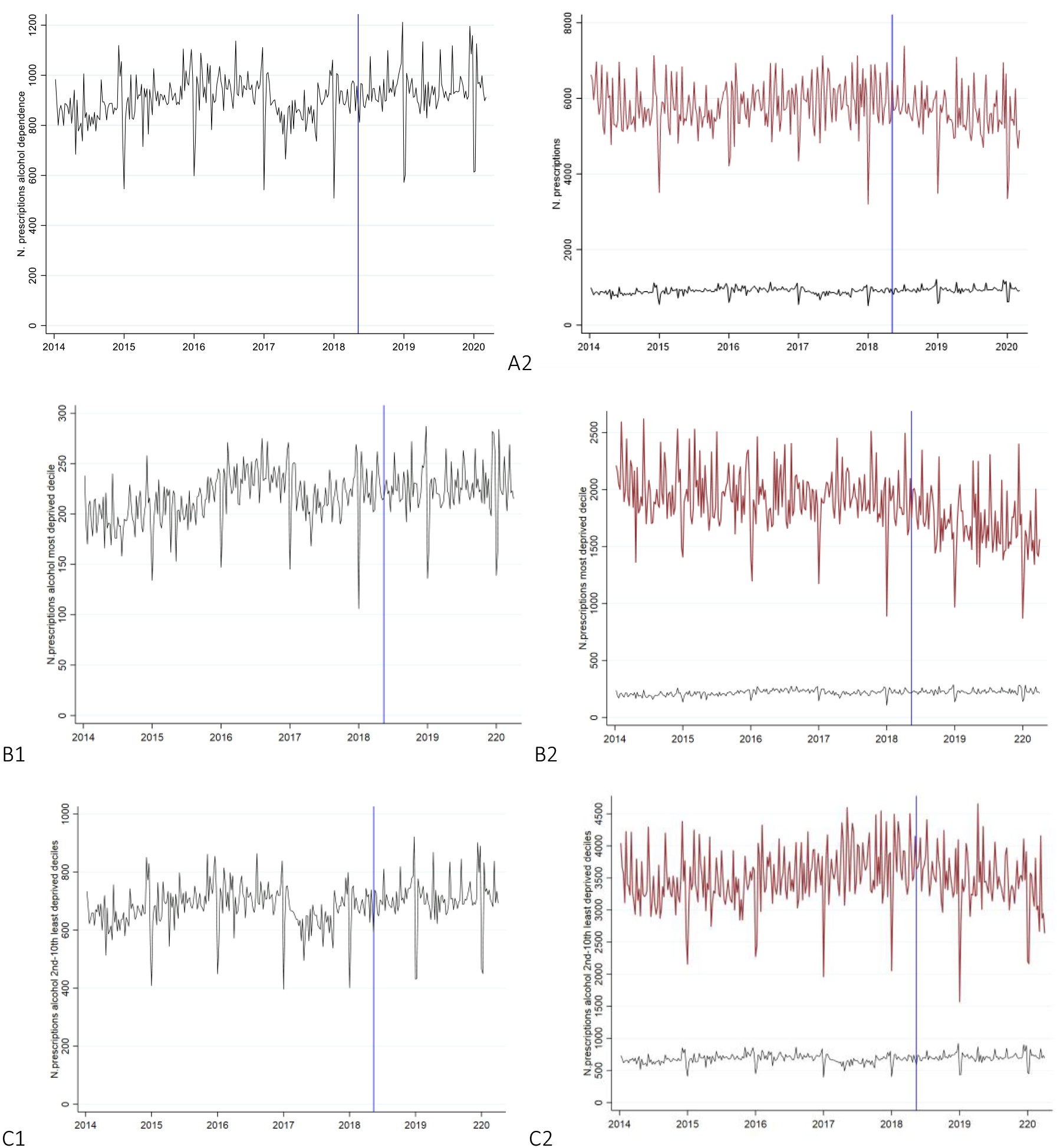
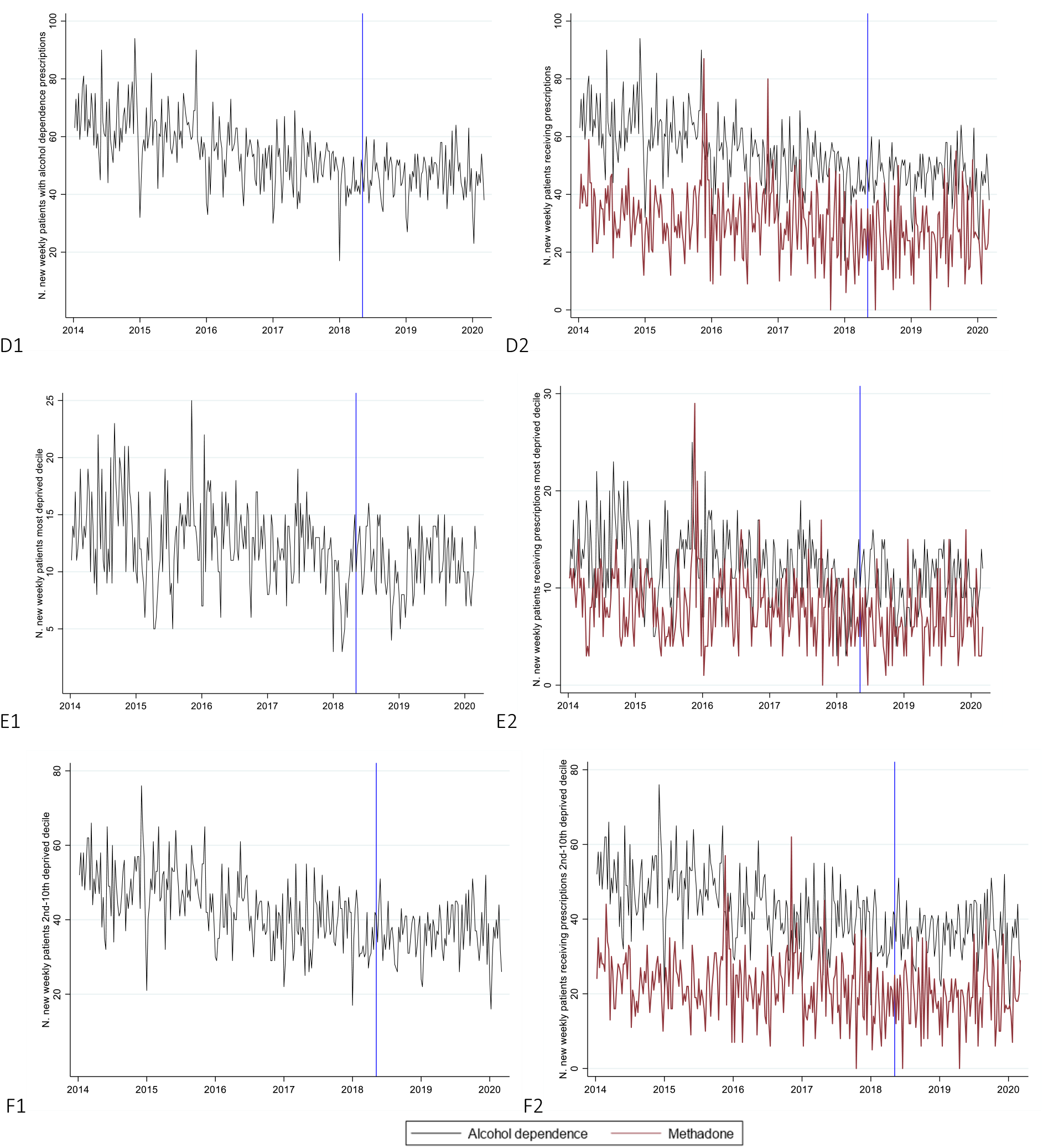
Trend in weekly prescriptions for alcohol dependence in Scotland (left column) and comparison between methadone and alcohol prescription in Scotland (right column) across different outcomes. A: Total number of prescriptions. B: Prescriptions in most deprived groups. C: Prescriptions in least deprived groups. D: New weekly patients receiving prescriptions. E: New weekly patients receiving prescriptions in most deprived groups. F: New weekly patients receiving prescriptions in least deprived groups.

#### Number of new weekly patients

The number of patients receiving prescriptions for treatment of alcohol dependence for the first time declined over the years (Table 1). The trend was stable between the most socio-economically deprived group (−22.4%) and the rest of the population (−23.7%). While there were major differences in the volume of prescriptions, new methadone patients followed a similar pattern: the overall decrease in patients was -21.3% and similarly distributed between socio-economic groups (−23.1% for the most deprived decile and -20.1% in the rest of the population).

### Inferential analysis

#### Prescriptions

For the overall population, MUP was associated with a non-statistically significant reduction in the change of level (−2.7%; 95%CI -0.068 0.014; p = 0.196) and the change in slope was estimated to be 0.0% (95%CI -0.001 0.000; p = 0.707) – Table 2. There were no significant variations corresponding with MUP introduction across different socio-economic groups. Methadone prescriptions showed a statistically significant decrease in the slope of trend after MUP of 0.1% (95%CI: -0.002 -0.001; p=0.000) in the overall population and in the subpopulations.

**Table 2.** Main Output for base case analysis and falsification tests for intervention and control groups

| Main analysis |  |  |  |  | Falsification tests |  |  |  |  |  |
| --- | --- | --- | --- | --- | --- | --- | --- | --- | --- | --- |
|  |  |  |  |  | 6-months pre MUP |  |  | 6-months post MUP |  |  |
| Model |  | Coefficient | 95% C.I. | P-value | Coefficient | 95% C.I. | P-value | Coefficient | 95% C.I. | P-value |
| Alcohol dependence |  |  |  |  |  |  |  |  |  |  |
| Prescriptions |  |  |  |  |  |  |  |  |  |  |
| Total population | MUP | -0.027 | (-0.068 0.014) | 0.196 | -0.870 | (-0.132 -0.042) | 0.000 | -0.013 | (-0.065 0.038) | 0.614 |
|  | after MUP trend | 0.000 | (-0.001 0.000) | 0.707 | 0.000 | (-0.001 0.000) | 0.126 | 0.000 | (-0.001 0.001) | 0.863 |
| Most deprived decile | MUP | -0.037 | (-0.125 0.051) | 0.408 | -0.118 | (-0.196 -0.040) | 0.003 | -0.019 | (-0.101 0.062) | 0.640 |
|  | after MUP trend | -0.001 | (-0.002 0.001) | 0.404 | -0.001 | (-0.002 -0.000) | 0.023 | -0.001 | (-0.002 0.001) | 0.479 |
| 2<sup>nd</sup>-10<sup>th</sup> deprived decile | MUP | -0.014 | (-0.062 0.035) | 0.582 | -0.084 | (-0.145 -0.023) | 0.007 | -0.005 | (-0.054 0.044) | 0.840 |
|  | after MUP trend | -0.000 | (-0.001 0.001) | 0.851 | -0.000 | (-0.001 0.000) | 0.298 | -0.000 | (-0.001 0.001) | 0.862 |
| New patients |  |  |  |  |  |  |  |  |  |  |
| Total population | MUP | 0.054 | (-0.022 0.129) | 0.165 | -0.008 | (-0.090 0.074) | 0.851 | 0.087 | (0.006 0.169) | 0.036 |
|  | after MUP trend | 0.002 | (0.001 0.003) | 0.002 | 0.002 | (0.001 0.003) | 0.000 | 0.001 | (-0.001 0.003) | 0.216 |
| Most deprived decile | MUP | -0.005 | (-0.218 0.208) | 0.964 | -0.222 | (0.410 -0.034) | 0.021 | -0.086 | (-0.314 0.142) | 0.462 |
|  | after MUP trend | 0.002 | (-0.001 0.006) | 0.182 | 0.002 | (-0.000 0.004) | 0.093 | 0.005 | (0.000 0.009) | 0.047 |
| 2<sup>nd</sup>-10<sup>th</sup> deprived decile | MUP | 0.041 | (-0.079 0.160) | 0.508 | 0.013 | (-0.099 0.126) | 0.816 | 0.083 | (-0.043 0.210) | 0.194 |
|  | after MUP trend | 0.002 | (0.000 0.003) | 0.027 | 0.002 | (0.001 0.003) | 0.001 | 0.001 | (-0.001 0.004) | 0.280 |
| Methadone |  |  |  |  |  |  |  |  |  |  |
| Prescriptions |  |  |  |  |  |  |  |  |  |  |
| Total population | MUP | -0.011 | (-0.036 0.015) | 0.410 | 0.020 | (-0.019 0.059) | 0.313 | -0.032 | (-0.57 -0.007) | 0.014 |
|  | after MUP trend | -0.001 | (-0.002 -0.001) | 0.000 | -0.001 | (-0.002 -0.001) | 0.000 | -0.001 | (-0.002 -0.000) | 0.009 |
| Most deprived decile | MUP | -0.006 | (-0.058 0.045) | 0.799 | -0.007 | (-0.068 0.055) | 0.829 | -0.023 | (-0.079 0.033) | 0.416 |
|  | after MUP trend | -0.001 | (-0.002 -0.000) | 0.002 | -0.001 | (-0.002 -0.001) | 0.001 | -0.001 | (-0.002 0.000) | 0.052 |
| 2<sup>nd</sup>-10<sup>th</sup> deprived decile | MUP | -0.030 | (-0.075 -0.015) | 0.195 | 0.010 | (-0.050 0.071) | 0.740 | -0.037 | (-0.088 0.015) | 0.163 |
|  | after MUP trend | -0.001 | (-0.002 -0.001) | 0.000 | -0.002 | (-0.002 -0.001) | 0.000 | -0.001 | (-0.002 0.000) | 0.009 |
| New patients* |  |  |  |  |  |  |  |  |  |  |
| Total population | MUP | -2.853 | (-6.198 0.492) | 0.095 | -3.600 | (-8.356 1.164) | 0.139 | -0.526 | (-3.300 2.248) | 0.71 |
|  | after MUP trend | 0.070 | (0.032 0.108) | 0.000 | 0.040 | (0.003 0.077) | 0.033 | 0.078 | (0.024 0.132) | 0.005 |
| Most deprived decile | MUP | -1.487 | (-3.072 0.099) | 0.066 | -0.653 | (-2.296 0.991) | 0.436 | 0.186 | (-1.447 1.820) | 0.823 |
|  | after MUP trend | 0.018 | (-0.010 0.045) | 0.208 | 0.002 | (-0.020 0.025) | 0.853 | 0.011 | (-0.027 0.049) | 0.579 |
| 2<sup>nd</sup>-10<sup>th</sup> deprived decile | MUP | -0.634 | (-4.868 3.599) | 0.769 | -0.602 | (-5.128 3.925) | 0.794 | -0.745 | (-4.131 2.641) | 0.666 |
|  | after MUP trend | -0.033 | (-0.035 0.102) | 0.341 | 0.018 | (-0.038 0.075) | 0.526 | 0.052 | (-0.027 0.131) | 0.195 |

\*\*number of new weekly patients receiving methadone had three value equal to 0 in the study period, therefore there was not log transformation of the dependent variable

#### New weekly patients

For weekly new patients, MUP was not associated with immediate changes. However, a significant increase in slope of the trend of 0.2% was estimated after MUP introduction for both the overall population and the least deprived deciles (Table 2). Regarding methadone, a significant gradual weekly decrease in the number of new patients was associated with MUP introduction.

### Sensitivity analysis

Whenever there were significant results in the intervention group, falsification tests bringing the analysed date of intervention forward by 6 months produced similar results (gradual change in new patients for overall population and least deprived groups). When an additional variable denoting the period post shortage of disulfiram was added to the regression in models with significant MUP terms, both this variable and the MUP variable became non-significant. When the MUP variable was removed and only the new ‘post shortage’ variable was left, this became significant and the model had a better fit (lower information criteria) (see supplementary material for these sensitivity analyses results).

The common trend assumption was satisfied only for volume of prescriptions for the total population, prescriptions in least socio-economically deprived groups, and most deprived new patients. When the ITS analysis was performed on the alcohol and methadone difference for such groups, we found significant associations only for slope change in prescriptions in the least deprived groups (−0.2% 95%CI: -0.003 -0.001; p=0.007). However, these differences were not robust to falsification testing.

## Discussion

This study provides evidence that MUP in Scotland was not associated with significant changes in the volumee, or trend in volume, of prescriptions for treatment of alcohol dependence in the overall population after 23 months of its introduction. For new patients receiving alcohol dependence medication, statistically important results were observed but this was mirrored in falsification tests anticipating MUP introduction by 6 months, thus making the effect unlikely to be causal. This suggests that time varying (unmeasured) confounders rather than the introduction of MUP led to an increase in the number of patients receiving such prescriptions for the first time.

We did not have any *a priori* hypothesis of the direction of a potential effect of prescriptions due to an introduction of a floor price for the purchasing of alcohol. Two plausible hypotheses are that the increase in price leads to decreasing consumption, resulting over time in decreased severity (or incidence) of alcohol dependence (possibly reducing demand for medications); conversely, the increase in price makes alcohol less affordable, increasing the likelihood that people with alcohol dependence may find alcohol unaffordable and be motivated to seek alcohol services (possibly increasing demand for medications). The latter is perhaps more likely in the time frame of our study. In any case, the null results we found could reflect that these two effects cancelled each other out, or that any effect was not large enough to be detected, or, that there is no causal effect of MUP intervention on level of uptake of alcohol dependence medications. Previous studies showing that people drinking alcohol at harmful levels did not change their alcohol consumption patterns due to MUP [18], as well as the similarity of trends with our control, provide support for the latter interpretation. Further, these findings may reflect limited capacity in alcohol treatment and a reluctance by some primary care doctors to prescribe treatments for dependence which may have been unchanged by MUP, even if more patients sought help[26]. Finally, another explanation for the null findings could be that the impact of MUPwas limited by inflation. The current £0.50 floor price was set in 2012, in 2021 the equivalent value would be almost £0.10 greater [29] (a 20% increase). The lack of an indexing price system risks to undermine even further the MUP effects (and potential benefits) in the future.While it has been shown that MUP reduced the overall alcohol consumption in the population [13], the demand for alcohol in alcohol dependent individuals can be less price elastic, explaining our results on both patients and prescriptions. In other words, a floor price of £0.50 (which did not affect all alcohol beverages on sale) was unlikely to decrease the real income and then holding back individuals from continuing/relapsing with alcohol dependence. This was also supported by recent evidence around MUP, showing that many of the people already drinking at harmful levels were not considerably affected by MUP as they already paid more than the floor price, with no substantial difference in consumption for those with alcohol dependence [18]. Are in line with this, and by showing that there was not a decrease in new patients receiving prescriptions after MUP, highlighted that the policy also was not effective to prevent people to be newly alcohol dependent (or at least did not change the number of new patients getting pharmaceutical treatment).

As mentioned, the few significant associations across subgroups and outcomes with MUP introduction can be attributed to time-varying confounding. Indeed, falsification tests registered significant changes for such subgroups where the ‘intervention’ was placed 6 months prior to the start of the MUP. In the model on the difference in prescriptions between alcohol and methadone, given the considerable initial difference in prescription levels, results may be led by methadone trends only. This would be supported by similar significant coefficients in the difference, falsification test of the difference (supplementary materials) and coefficient and falsification tests of the methadone analysis (Table 2). In addition, regarding new patients, when we inserted additional slope variables after the national shortage in disulfiram, they generated an attenuation (and lack of statistical significance) of the MUP effect. Furthermore, when such dummy variables were maintained, and MUP variables were excluded, the overall goodness of fit of the regression increased. From these tests, it is plausible to say that the significant effect found in the main analysis was likely caused by other factors happening earlier than MUP (if not the shortage in disulfiram).

The volume of prescriptions increased from 2014 to 2020. In contrast, the number of patients receiving prescriptions for alcohol dependence for the first time decreased over the same period. This discrepancy between the two series is difficult to explain; however, therapies can have long treatment periods or individuals can relapse, causing additional prescriptions but not additional patients. Regarding patients, a recent decrease in new patients in England receiving specialist alcohol treatment was found [30] which suggests that this fall was associated with financial pressures and service reconfiguration which prompted capacity reductions. Whilst our data are from community (primary care) prescribing, pressures on general practitioners’ capacity may have similarly strong effects on the number of patients in treatment and receiving prescriptions, perhaps more so than MUP.

### Strengths

To the best of our knowledge, this is the first time that a study has investigated changes in the level of prescriptions for treatment of alcohol dependence after the introduction of a MUP for alcohol. This is one of the few studies examining how MUP affected clinical outcomes associated with alcohol use/dependence which can inform the Scottish Government on the decision to extend or change the current policy, or allow it to lapse after the sunset clause deadline. Further studies with similar analyses on a longer time horizon could contribute to understanding of whether/how MUP may affect the rates and severity of alcohol dependence in the Scottish population over several years or decades.

We designed a population-based study, using data from whole of Scotland (and England) and thus removing any selection bias arising from sampling. Several robustness checks assessed the strength of our results.

### Limitations

England, being a neighbouring country and affected by the same shortage in disulfiram, could have been the ideal control group. However, there was a different prescription pattern between the two jurisdictions (Figure 1): disulfiram in England made up a considerably smaller proportion of total prescriptions for alcohol dependence treatments, limiting the relevance of the overall comparison with Scotland. Regarding the comparison with methadone prescriptions, we found that the two series had common pre-intervention trends for some analyses. Yet, our analyses using a difference-in-difference design did not find any effect of MUP after sensitivity analyses had been conducted. We recognise, however, that even with a common trend this may not have been the most reliable comparison, especially for prescriptions, having relevantly different initial volumes. The difference was smaller regarding patients. A further limitation is that we only accounted for methadone, while data on buprenorphine (the other drug with an indication for the management of opioid dependence in the UK, which is generally less prescribed [31]), was not available to us. Acknowledging that our controls presented a few weaknesses, we performed extensive sensitivity analysis on the uncontrolled series as well as on the controls, further validating our conclusions. Finally, many alcohol dependent individuals do not ever receive treatment medications [32, 33]: and new patients receiving such prescriptions are just a proportion of the new alcohol dependent individuals in the population, and medication may not be an appropriate treatment for all of them. Lastly, we recognise that dependence usually develops over a long period, and a longer analysis period could have helped to explain what might be happening. A challenge to modelling a longer time series is that after March 2020, the Covid-19 national lockdown considerably varied the prescription patterns. If we had extended analysis into this period it would have increased time-varying confounding.

## Conclusions

MUP was not associated with a change in the number of prescriptions for treatment of alcohol dependence in the overall population, neither with variations in the number of new patients receiving medication for alcohol dependence over a 23 months follow-up period. Further, there was no evidence of effect modification across different socio-economically deprived groups.

## Data Availability

Weekly data on Scotland are not publicly available due to disclosure reasons. Monthly data referring to England are available online at https://opendata.nhsbsa.net/dataset/english-prescribing-data-epd

## Author Contributions

Conceptualization, JL, FM; methodology, JL, FM; formal analysis, FM; data curation, FM, LZ; visualization, FM; supervision, JL, NF; interpretation, NF, JL, FM; writing – original draft preparation, FM; writing – reviewing and editing, NF, JL, DM, FM, AMc, CS, LZ; funding acquisition, not applicable. All authors have approved the final article.

## Acknowledgment

Data from the study were originally obtained from a separate study funded by Alcohol Research UK.

